# Serological Tests for SARS-CoV-2 Coronavirus by Commercially Available Point-of-Care and Laboratory Diagnostics in Pre-COVID-19 Samples in Japan

**DOI:** 10.1101/2020.07.10.20150904

**Authors:** Mariko Harada Sassa, Zhaoqing Lyu, Tomoko Fujitani, Kouji H. Harada

**Affiliations:** Department of Health and Environmental Sciences, Kyoto University Graduate School of Medicine, Yoshida Konoe, Sakyo, Kyoto 6068501, Japan

## Abstract

Serological tests for SARS-CoV-2 coronavirus in pre-COVID-19 samples in Japan showed 1.5%–1.75% positives, and previous surveys might overestimate COVID-19 seroprevalence in several population of Japan. These false negatives could be excluded by combination of different diagnostics to 0.25%.

## Text

The spread of the COVID-19 infection is still not fully understood. Serology is now employed to predict the cumulative incidence of COVID-19 infections *(1)*. The number of SARS-CoV-2 serological assays such as enzyme-linked immunosorbent assay (ELISA) and lateral flow assay (LFA) is increasing, but external validation has just started. Nonspecific reactions, cross-reactivity with various common pathogens including other human coronaviruses need to be evaluated *(2)*. In Japan, the number of COVID-19 patients is still small, but there may be more undiagnosed people. Some preprints and letters have demonstrated several percent seropositivity using LFA *(3,4)*. However, they were significantly higher than the number of PCR-determined symptomatic patients, and effects of false positives are suspected. Therefore, a cross-sectional study using specimens before 2019 from Japanese communities was conducted to evaluate the positive rate (non-specificity) by diagnostics available in Japan.

Cross-sectional serological survey of SARS-CoV-2 antibodies (Ig G and IgM) were performed on healthy adults in Kyoto Prefecture, Japan. Participants provided sera at local health check-ups with informed consents, and samples were archived in Kyoto University Environmental Biobank. 400 serum samples from 2012 to 2015 were used for this analysis (80 men, 320 women, average age 63.8 years, standard deviation 13.5 years). LFA (COVI040, GenBody Inc.) for detecting IgM or IgG against SARS-CoV-2 and ELISA for IgG (RAI010R, BioVendor Laboratories) were used. These diagnostics use the N protein of SARS-CoV-2 as an antigen. Weak signals in LFA were judged as positive. In the ELISA, the lower limit of signal intensity in COVID-19 patients, indicated by the manufacturer was used as the cutoff value. The analysis was initially conducted for 40 pooled samples from 400 serum samples (10 each by age and sex). Individual sera in pooled samples were examined for the positive group. All individual sera were considered negative if the pooled sera were negative. As a positive control, commercially available convalescent plasma of COVID-19 was used. To further verify reproducibility, individual IgG positive sera in the above LFA and ELISA were tested with another LFA (#CG-CoV-IgG, RayBiotech Inc.). All three diagnostics were for research use only and are not approved for clinical use in Japan. The Kyoto University Ethics Committee approved this study protocol.

Positive control samples were tested twice with three different test kits, and all showed positive results. In pre-COVID-19 samples, there were 6 IgG weak positives and 7 IgM weak positives among 40 pooled serum samples in LFA. The individual sera that made up the positive pooled samples were then examined, and 1 IgG positive, 5 IgG weak positives, and 7 IgM weak positives were observed (Table 1). By ELISA, 6 out of 40 pooled serum samples showed IgG positive, and 7 individual samples were IgG positive (1 pooled serum had 2 positives). Some other values were also seen near the cutoff value (Figure 1). The same tests were performed again on the above positive samples, and a positive reaction was confirmed. From the results, the false positive rate was 1.5% for IgG (95%CI: 0.552%–3.24%), 1.75% for IgM (95%CI: 0.706%–3.57%) in LFA. In ELISA, it was estimated to be 1.75% (95% CI: 0.706%–3.57%). These positive cases were partially overlapped between tests. Only one case showed positive for both IgG and IgM in LFA. Another one patient showed positive by both LFA and ELISA (0.25%; 95%CI: 0.006%–1.39%) (Table 1). 12 IgG positive samples were further assayed with additional LFA, and weak signals were detected in 1 of 6 LFA IgG positives and 2 of 7 ELISA IgG positives. None of the samples tested positive in all three tests.

**Table 1.** Comparison of anti-SARS-CoV-2-NP IgG and IgM assay specificities in LFA and ELISA in Pre-COVID-19 sera.

| (A) Test results for IgG between LFA and ELISA |  |  |  |
| --- | --- | --- | --- |
| no. (%) | LFA (+) | LFA (-) | Total |
| ELISA(+) | 1<br>(0.25%) | 6 (1.5%) | 7 (1.75%) |
| ELISA(-) | 5<br>(1.25%) | 388 (97%) | 393 (98.25%) |
| Total | 6 (1.5%) | 394 (98.5%) | 400 |
| (B) Test results between IgG and IgM in LFA |  |  |  |
| no. (%) | IgG(+) | IgG(-) | Total |
| IgM(+) | 1<br>(0.25%) | 6 (1.5%) | 7 (1.75%) |
| IgM(-) | 5<br>(1.25%) | 388 (97%) | 393 (98.25%) |
| Total | 6 (1.5%) | 394 (98.5%) | 400 |

LFA: lateral flow assay, ELISA: enzyme-linked immunosorbent assay, (+): positive, (-): negative.

**Figure 1.**
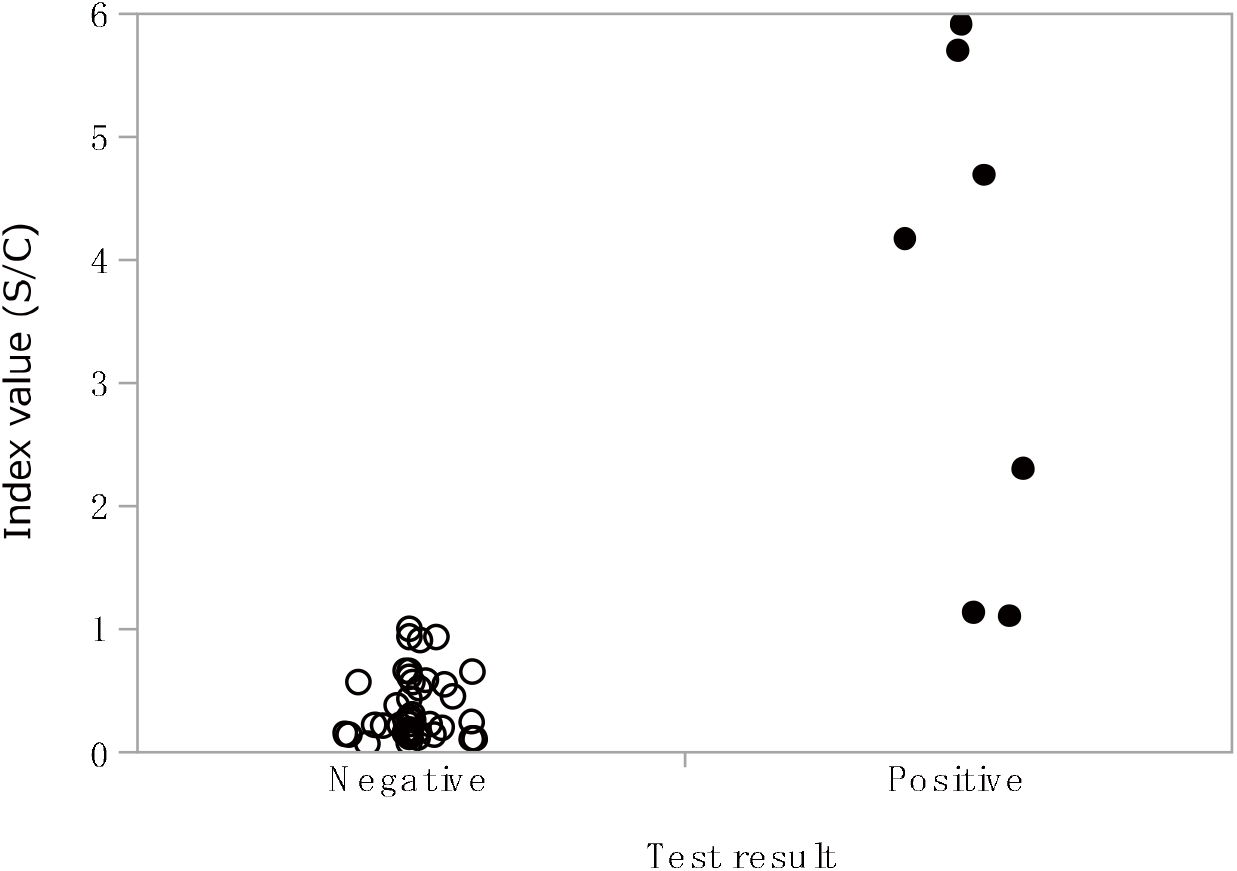
Distribution of signals detected by anti-SARS-CoV-2 NP ELISA in Pre-COVID-19 sera. Index value, Signal to cut-off (S/C) ratios are plotted.

This study examined the nonspecific reactions of serological testing using pre-COVID-19 sera in Japan. Both LFA and ELISA used in this study showed a positive reaction over 1%. The IgM test was usually more nonspecific than IgG test, but IgG also had similar false positive rates. It is likely difficult to estimate the prevalence of COVID-19 by antibody testing in areas where COVID-19 is not prevalent at high rates. In Japan, the number of confirmed COVID-19 infections was 19,282 until 4 Jul, 2020 *(5)*, which is 0.015% of the population of Japan, 125 million. Even when the true seroprevalence of the population were 0.1% and asymptomatic individuals were tested by an assay with 95% sensitivity and 98.25% specificity, the false discovery rate would be approximately 95%. Reported IgG seroprevalence in several populations in Japan showed around 3–5%. However, given that the false positive rate in this study can be applied, those numbers would consist of nonspecific reactions at a high rate. Specificities also vary among researches and diagnostics due to qualitative readings, cutoff values, etc. Nevertheless, in this study, there was almost no agreements in different test kits for false positives, then it is possible to reduce the false positives by using multiple diagnostics for positive samples.

## Data Availability

All data is included in the preprint.

## Acknowledgments

The authors sincerely express their appreciation to contributors for the Kyoto University Environmental Biobank.

This study was supported by a Grant-in-Aid for Scientific Research from the Japan Society for the Promotion of Science. The funder had no role in this study. We have no conflicts of interest.

## Notes

### Competing Interest Statement

The authors have declared no competing interest.

### Author Declarations

The Kyoto University Ethics Committee approved this study protocol.

